# A series of pulses vaccination in SIR model – Understanding periodic orbits and irregular trajectories

**DOI:** 10.1101/2022.05.30.22275782

**Authors:** Hyun Mo Yang

## Abstract

When a Susceptible-Infective-Recovered (SIR) model with a constant contact rate is used to describe the dynamics of directly transmitted infections, oscillations, which decay exponentially with time, are obtained. Due to damped oscillations, intermittent vaccination schemes can be designed in order to reduce or even eliminate the infection. A simple intermittent vaccination can be described by a series of pulses, i.e., a proportion of susceptible individuals is vaccinated intermittently at every fixed period of time. Analysis of the model is done by numerical simulations in order to determine the trajectories in the phase space. It is observed that as the proportion of vaccinated individuals increases, closed orbits with multiple cycles appear, even irregular trajectories arise occasionally. These results can be understood by comparing with bifurcations occurring in a discrete logistic model describing a single population. Further, bifurcations occurring in epidemiological models that use periodic functions to mimic seasonal variations in the disease transmission are discussed.

## 1 INTRODUCTION

Directly transmitted infections are mathematically modelled by assuming a homogeneous mixing among them, that is, individuals interact randomly at a certain rate. The classical SIR (Susceptible–Infective–Recovered) type of model considering a constant contact rate yields oscillations which decay exponentially, *i*.*e*., damped oscillations [2]. In [23], by introducing a pulse perturbation in an SIR model, the natural-epidemics (time lag between two consecutive peaks of the natural epidemic) and the inter-epidemics (time lag between two consecutive peaks of the perturbed disease transmission system) periods were estimated. However, when time-dependent model parameters are considered, the natural-epidemics period disappears, and annual cycle epidemics arises (see [25] for a dengue modelling).

The oscillating patterns in the dynamics of directly transmitted infections allow the use of an intermittent vaccination strategy, together with or instead of a routine scheme. The reason behind it is the application of successive mass vaccinations always just before the triggering of the epidemics. Mass vaccinations in a short period of time can be roughly described by a series of pulses vaccination, that is, a strategy where a pre-determined proportion of susceptible individuals is vaccinated intermittently at a fixed time intervals. The time between successive pulses could be determined by the natural-epidemics period. For instance, according to data published by the Centers for Disease Control, the basic pattern of the rubella epidemics is a 3-*year* cycle. However, the epidemics cycles from 6-to 9-*year* intervals present a higher incidence as a result of a buildup and fall in incidence over the basic cycle [9].

In developed countries the routine approach is applied with very good results [8] due to well organized health systems. This is not true, however, for developing countries where an alternative compounded strategy is applied, *i*.*e*., routine vaccination associated with complementary mass vaccination campaigns. The reasons for applying this compounded strategy are derived from the lack of organization and continuous resources for the health system, conjugated with the lower population compliance to routine programmes. The experience in Brazil with this approach has been very encouraging for poliomyelitis [22], measles [17] and more recently with rubella [12].

Can regularly applied pulses vaccination arise complex dynamics? In order to address this question, numerical analysis of SIR model encompassing pulses vaccination strategy is done. As the proportion of susceptible individuals is increased, closed orbits with multiple cycles arise, with irregular trajectories appearing occasionally. A discrete logistic model [13] shed some light to interpret these results. Even more complex behavior appearing in SIR model with periodic (sinusoidal) function [20] can be understood. Another question addressed here is the assessment of the minimum proportion of susceptible individuals under vaccination such that the disease can be considered eradicated.

The paper is divided as follows. In section 2, a vaccination strategy comprised in a series of pulses is proposed based on the SIR model. In section 3, scenarios for different proportion of vaccinations are shown, and discussion is given in section 4. Conclusion is given in section 5.

## 2 SIR MODEL

Periodicity of some infectious diseases, especially childhood infections like measles and rubella, has attracted attention of many researchers. Although not fully understood, it is very important to take it into account when designing an immunization programme.

Dynamics of directly transmitted infections, disregarding maternally derived antibodies and differential mortality due to the disease, is considered. In the modelling, a life-long immunity, induced by both the natural infection and the vaccine, is assumed. A closed community is divided into three classes named susceptible, infectious and recovered individuals, with the fractions of respective classes at time *t* being denoted by *s*(*t*), *i*(*t*) and *r*(*t*). Instead of infectious individuals *i*(*t*), the force of infection *λ*(*t*), defined by

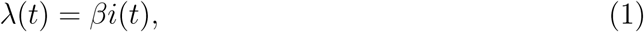

is taken into account in the modelling. The parameter *β* is the transmission coefficient being given by *β* = *β*′*N*, where *β*′ is the constant per-capita contact rate and *N* is the constant population size.

Assuming that the susceptible and infectious individuals are mixed homogeneously in the community [3], the SIR model is, then, described by

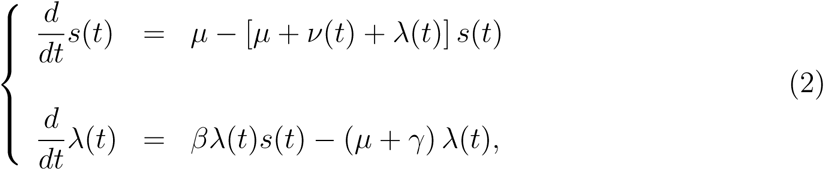

where *r*(*t*) = 1 − *s*(*t*) − *λ*(*t*)*/β*, which comes out from the assumption that we are dealing with a constant population size, that is, the birth rate is equal to the natural death rate, denoted by *µ*. The parameter *γ*^−1^ is the average recovery period (*γ* is the recovery rate) and *ν*(*t*) is the vaccination rate applied to susceptible individuals in all ages.

The analysis of the model given by equation (2) with constant *ν* was done in [24]. Briefly, suppose that *ν*(*t*) = 0. Equation (7), without vaccination, has two equilibrium points denoted by 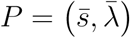 (see [23] for details): one is the trivial, given by *P* ^0^ = (1, 0), and the other is the non-trivial *P*^*^ = (*s*_0_, *λ*_0_) with coordinates

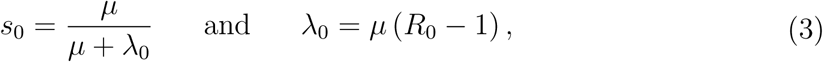

where the basic reproduction number *R*_0_ is

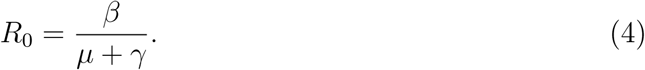

When a constant and continuous vaccination *ν* is applied on a population (with *R*_0_ *>* 1), there is a reproduction number due to the vaccination *R*_*ν*_, given by *R*_*ν*_ = *µR*_0_*/* (*µ* + *ν*), such that *R*_*ν*_ = *R*_0_, for *ν* = 0, and *R*_*ν*_ *< R*_0_, for *ν >* 0 [2]. Clearly there is a threshold of *ν*, denoted by *ν*^*th*^, given by

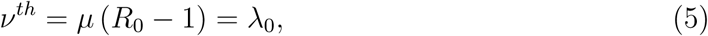

which, for all *ν > ν*^*th*^, the disease is eradicated by vaccination.

The application of mass immunization strategy consisting of a series of pulses as a vaccination scheme is proposed and evaluated. This type of vaccination arises the question of the choice of the time interval between two successive pulses immunization. The main aspect about this question is related to the outbreak of severe epidemics when susceptible individuals prevail in the population. For example, in a long time delayed pulses vaccination scheme, the number of susceptible individuals who were not covered by each successive immunizations is summed up and increases, and, consequently, a small number of infectious individuals can trigger a severe epidemics [23]. To avoid this undesirable effect, the time lag between successive pulses must be chosen appropriately, for instance, the natural-epidemics period multiplied by a factor.

Based on the above reasonings, a mass vaccination in the form of pulses [1] is described by the vaccination rate *ν*(*t*) given by

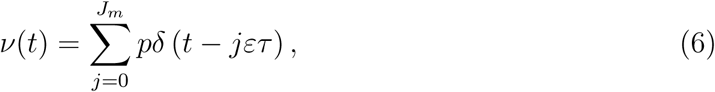

where *J*_*m*_ is the number of pulses vaccination applied, and *δ*(*t*) is the Dirac delta function [4]. The parameters *τ* and *ε* are respectively the natural-epidemics period and security factor, with preferentially *ε <* 1, and *p* is the effective proportion of the susceptible individuals covered by vaccination at time *t*_*j*_ = *jετ*. Hethcote [10] used this form of vaccination rate to determine the optimal vaccination age.

A series of pulses vaccination strategy can be described by equation (2) dropping out *ν*(*t*), that is,

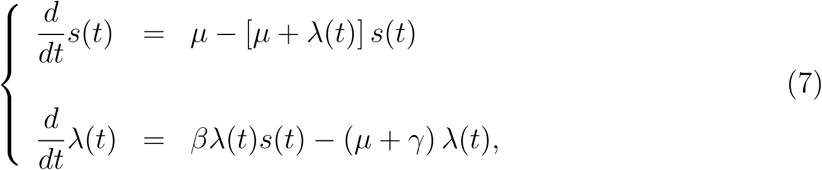

and the Dirac delta pulses are transferred to boundary conditions, which are

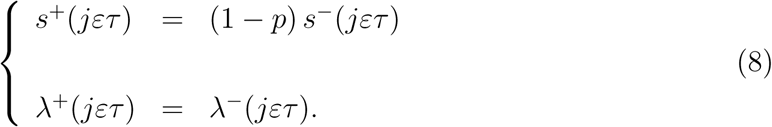

The superscript ^+^ stands for the limit of time coming from the right hand (*t*_*j*_ ← *t*) and ^−^, for the limit coming from the left (*t* → *t*_*j*_). Notice that at *t* = 0, the initial conditions are *s*^+^(0) = (1 − *p*) *s*_0_ and *λ*^+^(0) = *λ*_0_, where *s*_0_ and *λ*_0_ are given by equation (3).

By varying the proportion of susceptible individuals covered by the immunization, the scenarios yielded form application of a series of pulses vaccination strategy are described.

## 3 Studying pulses of vaccination

A series of pulses vaccination strategy, described by the vaccination rate given by equation (6), is based on three parameters: the natural-epidemics period *τ*, the security factor *ε* and the proportion covered by the vaccination scheme *p*. Here *τ* and *ε* are fixed, and only *p* is allowed to vary. Rubella infection is taken as an example.

Numerical simulations of the system of equations (7) by supplying the boundary conditions (8) are done, in order to obtain different scenarios (or, bifurcations). Numerical solutions are obtained by applying the stepsize controlled fourth order Runge-Kutta method [18]. Figures are shown in the phase space portrait *λ*×*s* containing the non-trivial equilibrium *P*^*^ given by equation (3).

Besides the evaluation of pulses as a vaccination scheme, another goal is the understanding of the complex dynamical trajectories. Since the time between successive pulses is fixed (*ετ*) but the proportion *p* is varied, periodic impulsive system with varying amplitudes is considered here. This study can be useful to understanding results of modelling the seasonal variations mimicked by periodic sinusoidal functions.

### 3.1 Model parameters

Using the field data from the City of Caieiras, São Paulo State, Brazil [7], the steady state force of infection was estimated for Rubella, which was *λ*_0_ = 0.0766 *years*^−1^. Assuming that *µ* = 0.016 *years*^−1^, the steady state fraction of susceptible individuals is *s*_0_ = 0.1728, and the basic reproduction number is *R*_0_ = 5.787, from equation (3). Finally, letting *γ* = 30.4 *years*^−1^, the transmission coefficient is *β* = 176.03 *years*^−1^ from equation (4). For rubella infection, the natural-epidemics period is *τ* = 4.118 *years* [23].

The quasi-periodic trajectories of the SIR model are illustrated in Figure 1, assuming that few infectious individuals are introduced in a susceptible population. Equation (2) is simulated assuming that there is not vaccination (*ν* = 0), with initial conditions given by *s*(0) = 1 and *λ*(0) = 10^−5^ *years*^−1^, or *i*(0) = *λ*(0)*/β* = 57 × 10^−9^ (57 individuals in 1 billion). After huge amplitudes in the beginning (Figure 1(a), shown until 30 *years*), oscillations are damped toward the non-trivial equilibrium *P*^*^ = (*s*_0_, *λ*_0_) = (0.1728, 0.0766*years*^−1^) (filled square), which is shown inside of the damped oscillations (Figure 1(b), shown from 100 to 150 *years*). The first peak of epidemics occurs at *t*_1_ = 0.15 *years*, with *λ*(*t*_1_) = 88, 71 *years*^−1^ (*i*(*t*_1_) = 0.504), and the next second peak occurs at *t*_2_ = 21.39 *years*, with *λ*(*t*_2_) = 4.80 *years*^−1^ (*i*(*t*_2_) = 0.027). Hence, the first inter-epidemics period is *θ* = *t*_2_ − *t*_1_ = 21.24 *years*.

**Figure 1:**
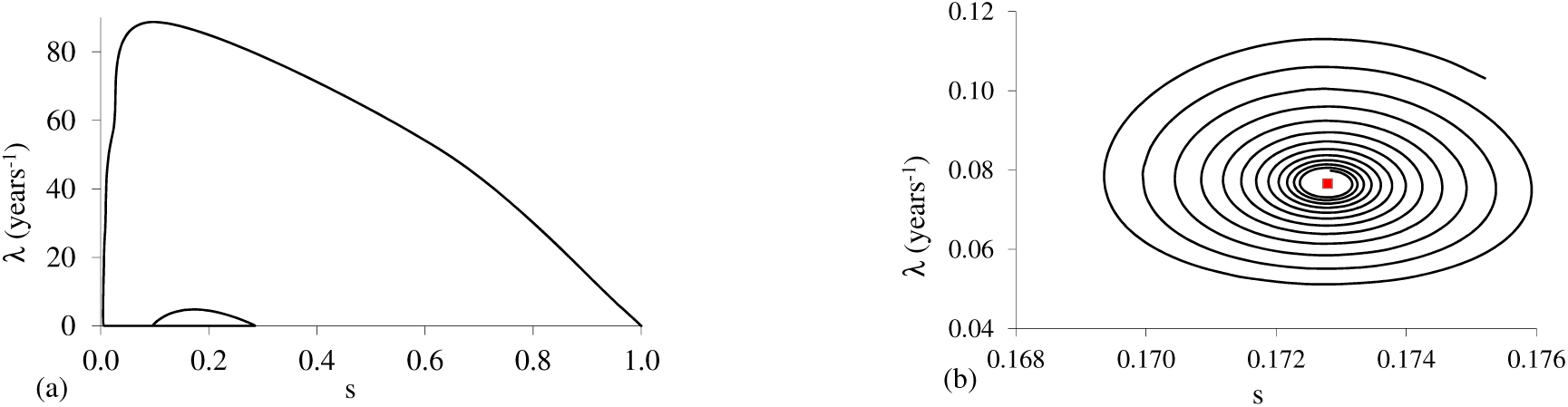
Introduction of an infection in a community free of disease without vaccination (*ν* = 0), described by the initial conditions *s*(0) = 1 and *λ*(0) = 10^−5^ *years*^−1^: phase portrait of (a) initial time (until 30 *years*), and (b) intermediate time (from 100 to 150 *years*), showing damped oscillations toward the non-trivial equilibrium *P*^*^ (filled square).

Figure 2 illustrates the time varying fraction of susceptible individuals (*s*) and the force of infection (*λ*) corresponding to Figure 1 disregarding the initial huge amplitudes (shown from 40 to 100 *years*). Clearly, the time elapsed between two consecutive peaks of epidemics decreases, and as *t* increases, the period of time between successive epidemics tends to the natural-epidemic period *τ* (see [23]).

**Figure 2:**
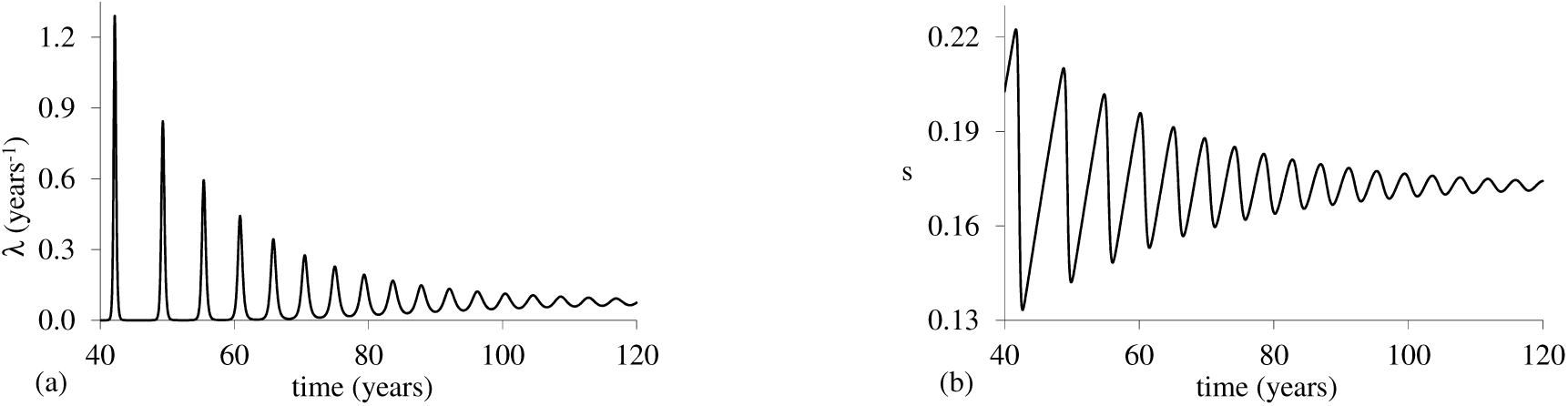
The time varying fraction of susceptible individuals (*s*) and the force of infection (*λ*) from 40 to 100 *years*, corresponding to Figure 1.

### 3.2 Evaluating pulses vaccination strategy

In an attempt to avoid the emerging of severe epidemics, a mass vaccination described by a series of pulses is applied in a community with equal period of time between successive vaccinations, being this period lower than the nature-epidemics period given by *ετ*.

When *p* = 0, damped oscillations drive the dynamical trajectories to attain the non-trivial equilibrium point *P*^*^ (see Figures 1 and 2). For *p* ≠ 0, periodic orbits must arise due to the periodically forcing external input (pulses vaccination with period *T* = *ετ*), which must be preceded by initial fluctuations due to transient effect. The period *T* of closed orbit is obtained solving numerically equation (7) by supplying the boundary conditions (8) as follows: (1) after elapsed a sufficient time until the transient effects fade out, at an arbitrary time *t*_1_ the values *s*(*t*_1_) and *λ*(*t*_1_) are stored, (2) for *t > t*_1_, the values *s*(*t*) and *λ*(*t*) are compared with *s*(*t*_1_) and *λ*(*t*_1_), (3) if inequalities defined by

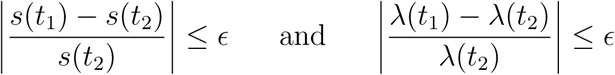

are satisfied, then *s*(*t*_1_) = *s*(*t*_2_) and *λ*(*t*_1_) = *λ*(*t*_2_) are assumed and, then, (4) the period of closed orbit is calculated by *T* = *t*_2_ − *t*_1_. Another periods are also searched. For the relative error, it is considered *ϵ* = 10^−6^.

All simulations are obtained considering security factor being *ε* = 0.8 and by varying the proportion of susceptible individuals vaccinated *p*. Hence, the following scenarios are related to a community submitted to a series of pulses with the time lag of *T* = *ετ* = 3.2944 *years* between them. The number of pulses vaccination performed, *J*_*m*_, is set in such a way that the vaccination is applied over all the time of simulation.

Figure 3 shows the case *p* = 5%: (a) initial time (until 50 *years*), and (b) long time (from 100 to 300 *years*). The closed orbit contains the non-trivial equilibrium *P*^*^, and this orbit has the same period as the periodic pulses vaccination, that is, *T* = *ετ* = 3.2944 *years* [21]. The non-trivial equilibrium *P*^*^ is shown as a filled square and is presented in all figures hereafter.

**Figure 3:**
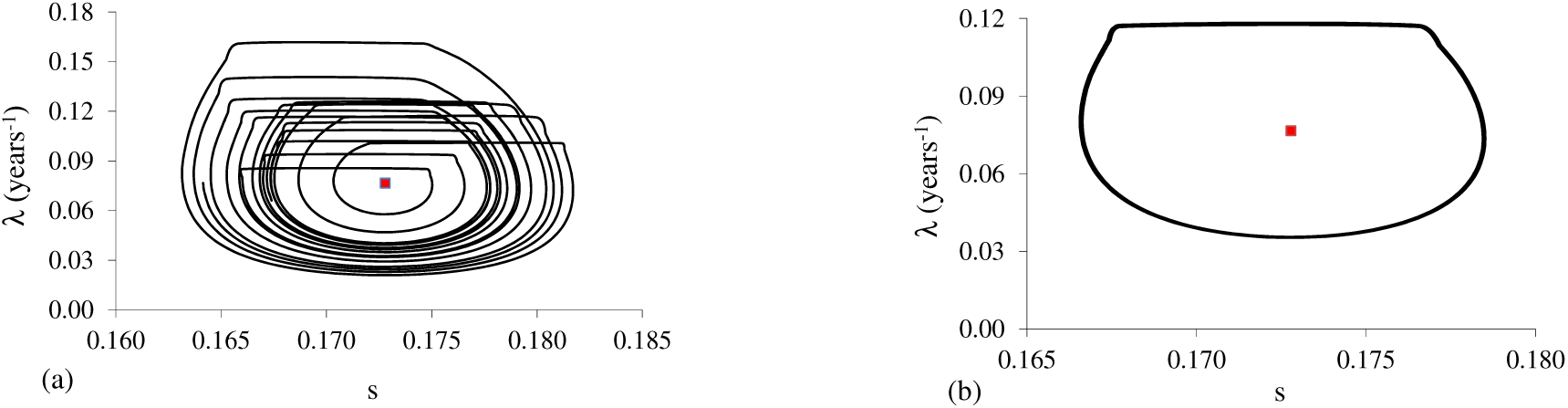
Phase space portrait for *p* = 5%: (a) initial time (until 50 *years*) and (b) long time (from 100 to 300 *years*), a one cycle orbit.

The closed orbits are formed after the transient period, which can last for more than hundreds of years as *p* increases. For this reason, the periodic orbits are searched after elapsing more than 160, 000 *years*, which is an arbitrary but sufficiently higher value to fade out transient behavior. The asymptotic phase space trajectories are shown from 163, 500 to 164, 000 *years*, which are called asymptotic curves.

As shown in Figure 3, for 0 *< p* ≤ 8.987%, damped oscillations are driven to a single closed orbit (limit cycle), with period *T* = *ετ*, that is, one cycle orbit persists until *p* = 8.987%. As *p* increases, *λ* and *s* circulate around *P*^*^, but with increased amplitudes.

For *p >* 8.987%, closed orbits whose period is multiple of *ετ* appear, besides irregular trajectories. At *p* = 8.987% occurs a flip bifurcation.

Figure 4 shows the case *p* = 8.988%: (a) initial time (until 50 *years*), and (b) asymptotic curve. The period of closed orbit is *T* = 2*ετ* = 6.5888 *years*. At this vaccination coverage occurs closed orbit of two cycles and doubling of the period.

**Figure 4:**
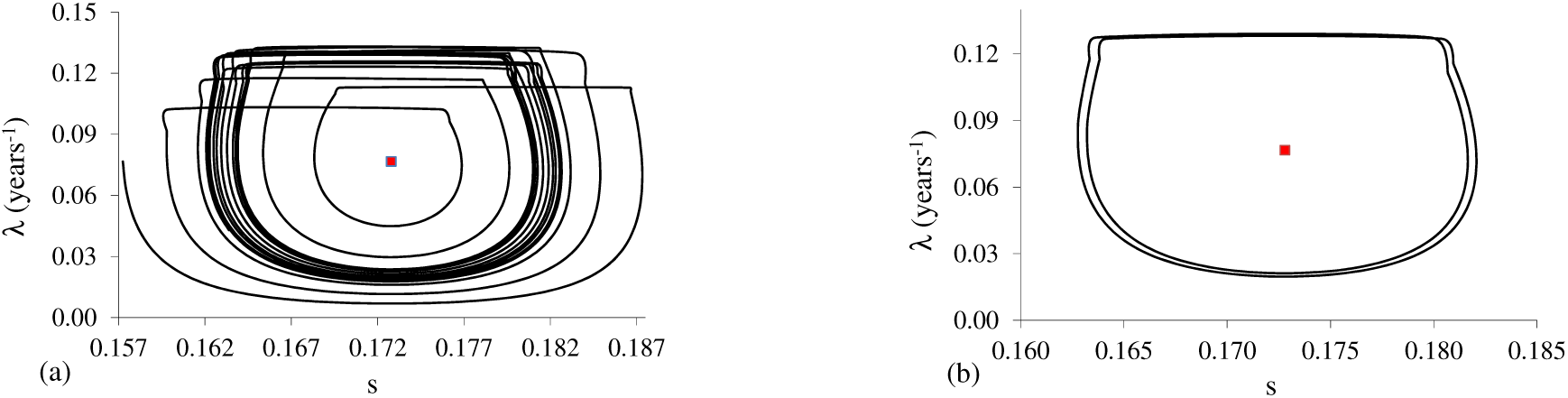
Phase space portrait for *p* = 8.988%: (a) initial time (until 50 *years*) and (b) asymptotic curve, a two cycles orbit.

Two cycles orbit persists until *p* = 9.316%. It is worth to showing behavior for intermediate and upper bound values of *p*. Figure 5 shows the asymptotic curves for: (a) *p* = 9.1%, and (b) *p* = 9.316%. Figures 4(b), 5(a) and 5(b) show clearly that the difference between the maximum values of *s* in both cycles increases as *p* increases.

**Figure 5:**
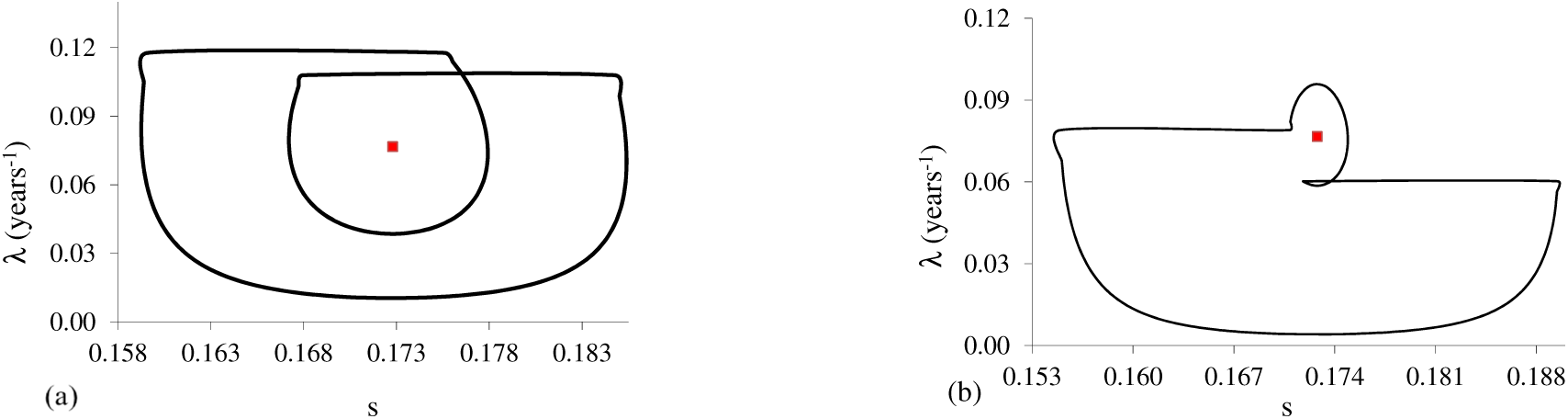
Asymptotic two cycles phase space portrait for: (a) *p* = 9.1% and (b) *p* = 9.316%.

The origin of the two cycles orbit is due to the enlargement in the amplitude of oscillation of the fractions of susceptibles (*s*) by increasing *p* (vaccination of susceptible individuals). Figure 4(b) shows practically coincident orbits, but as *p* increases, the increasing in the amplitude of *s* in one of two cycles orbit decreases the amplitude of the second cycle (internal orbit, Figure 5). The effect on *λ* as *p* increases is the splitting of two cycles, one with high amplitude, and the other with low amplitude. However, the closed orbit of two cycles could be destabilized by increasing *p*. This behavior has correspondence in a simple logistic discrete model [13], which is discussed below.

Figure 6 shows the case *p* = 9.317%: (a) initial time (until 50 *years*), and (b) asymptotic curve. The are not closed orbits, and the behavior is irregular, but circulating the equilibrium point.

**Figure 6:**
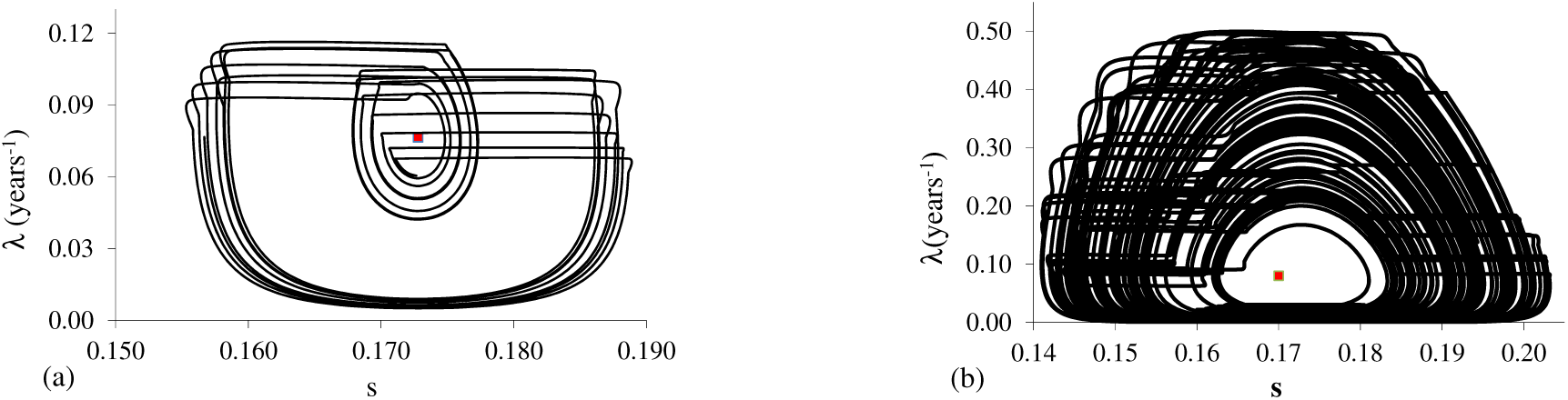
Phase space portrait for *p* = 9.317%: (a) initial time (until 50 *years*) and (b) asymptotic curve, an irregular trajectory.

Irregular behavior persists until *p* = 15.24%. To explore this irregular behavior, Figure 7 shows the asymptotic curves for: (a) *p* = 15.23%, and (b) *p* = 15.24%.

**Figure 7:**
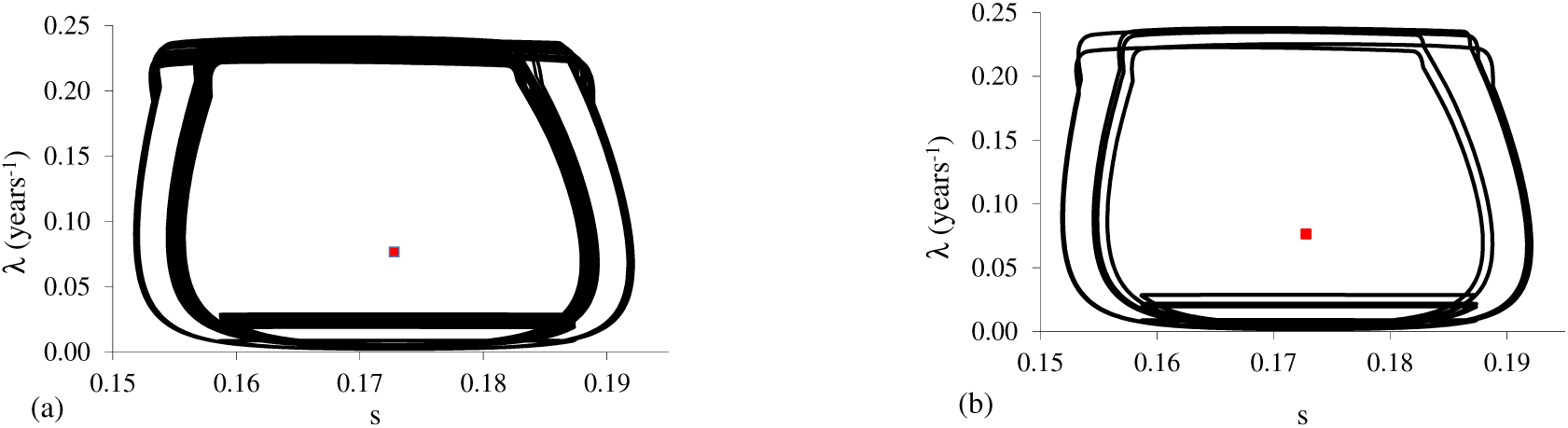
Asymptotic irregular phase space portrait for: (a) *p* = 15.23% and (b) *p* = 15.24%. Decreasing the relative error *ϵ*, periodic orbits are found.

For *p* = 15.22% (not shown here), there is a shrinking of trajectories observed in Figure 6(b): after large perturbations, the phase space trajectory is settle in a quasi rectangular box. Figure 7(a), for *p* = 15.23%, shows that the trajectories are bounded in a strip, and closed orbits are not observed (this is true for error *ϵ* ≤ 4.94 × 10^−4^). Figure 7 can be classified as torus bifurcation. However, if we decrease the accuracy (error is increased) to *ϵ* ≥ 4.95 × 10^−4^, then the orbit can be classified as closed with *T* = 308*ετ* = 1, 014.68 *years*, that is, the period is increased in 308 times. In Figure 7(b), for *p* = 15.24%, the strip is reduced, but there are not closed orbits for *ϵ* ≤ 7.29543 × 10^−5^. If the error is increased to *ϵ* ≥ 7.29544 × 10^−5^, then two closed orbits, with periods *T*_1_ = 56*ετ* = 184.486 *years* and *T*_2_ = 1776*ετ* = 5, 850.85 *years* are found. Figure 7 shows a trend for higher *p*: two types of cycles, where one has low amplitude in *λ*, and the other, high amplitude cycling the equilibrium point *P*^*^.

As *p* increases from 9.317%, the irregular behavior is changed to bounded trajectories. Hence, it is expected that increasing *p*, closed orbits could be achieved. As *p* varies since 15.25%, closed orbits appear, with periods decreasing until *p* = 17.437%, in which case a close orbit with periodicity *ετ* occurs.

Figure 8 shows the case *p* = 15.25%: (a) initial time (until 50 *years*), and (b) asymptotic curve. The period of closed orbit is *T* = 16*ετ* = 52.7104 *years*, and this sixteen cycles orbit persists until *p* = 15.26%.

**Figure 8:**
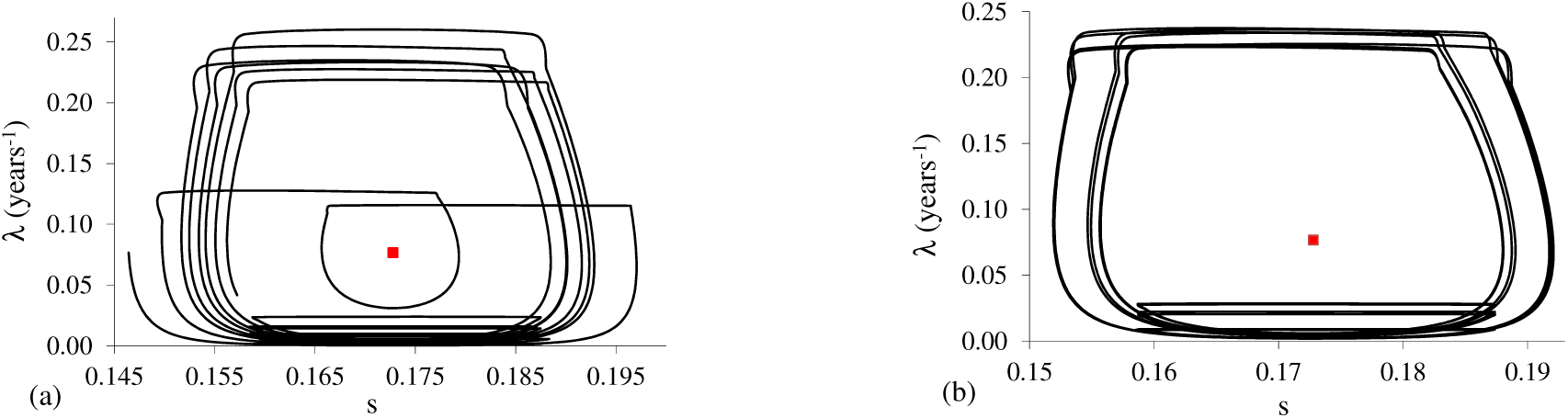
Phase space portrait for *p* = 15.25%: (a) initial time (until 50 *years*) and (b) asymptotic curve, a sixteen cycles orbit.

Figure 9 shows asymptotic curves for two values of *p*. When *p* = 15.27% (a), the period of closed orbit is *T* = 8*ετ* = 26.3552 *years*, and eight cycles orbit persists until *p* = 15.32%. When *p* = 15.33% (figure not shown), the period of closed orbit is *T* = 4*ετ* = 13.1776 *years*, which persists until *p* = 15.70%. When *p* = 15.71% (figure not shown), the period of closed orbit is *T* = 2*ετ* persisting until *p* = 17.436%. When *p* = 17.437% (b), a one cycle orbit is observed with period *T* = *ετ*, which persists until *p* = 18.926%. Notice that the closed orbit is formed with the equilibrium point *P*^*^ situating near the boundary of the line segment joining the points before and just after the pulse vaccination.

**Figure 9:**
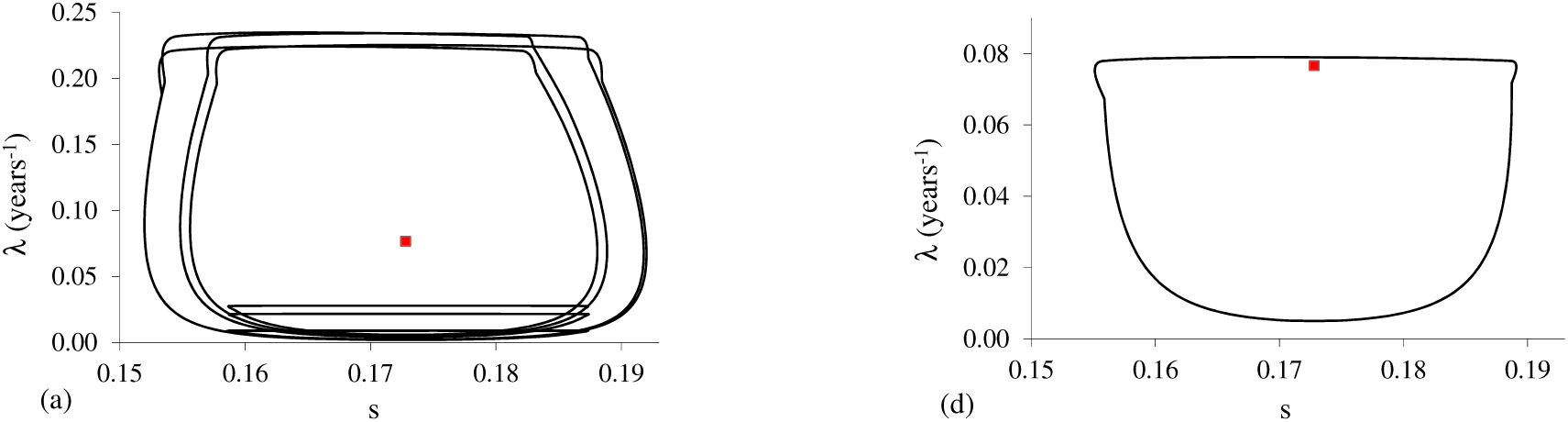
Phase space portrait for: (a) *p* = 15.27% with eight cycles, and (b) *p* = 17.437% with one cycle.

Figure 10 shows the case *p* = 18.927%: (a) initial time (until 50 *years*), and (b) asymptotic curve. The period of closed orbit is *T* = 6*ετ* = 19.7664 *years*. Six cycles orbit persists until *p* = 18.928%.

**Figure 10:**
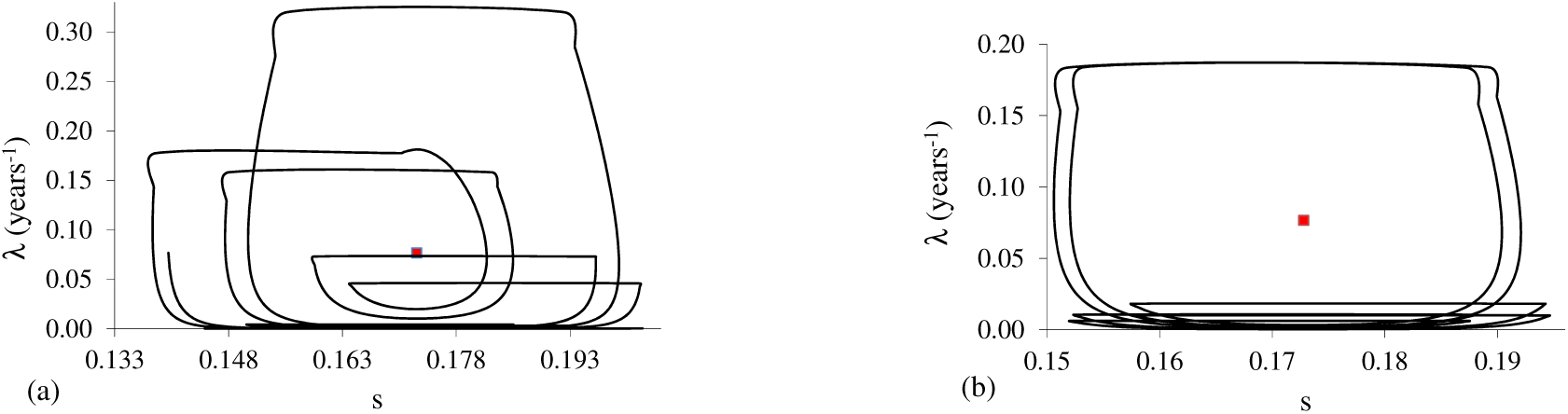
Phase space portrait for *p* = 18.927%: (a) initial time (until 50 *years*) and (b) asymptotic curve, a six cycles orbit.

There are alternating one cycle – six cycles orbits (11 alterations were observed between 18.929% and 18.974% by varying *p* in 0.001%; could other alternations be found if the increment is lower than 0.001%ν). The last six cycles orbit occurs between 18.973% and 18.974%. Figure 11 shows the asymptotic curves for: (a) *p* = 18.972%, and (b) *p* = 18.974%. The periods of closed orbits are *T* = *ετ* and *T* = 6*ετ*, respectively.

**Figure 11:**
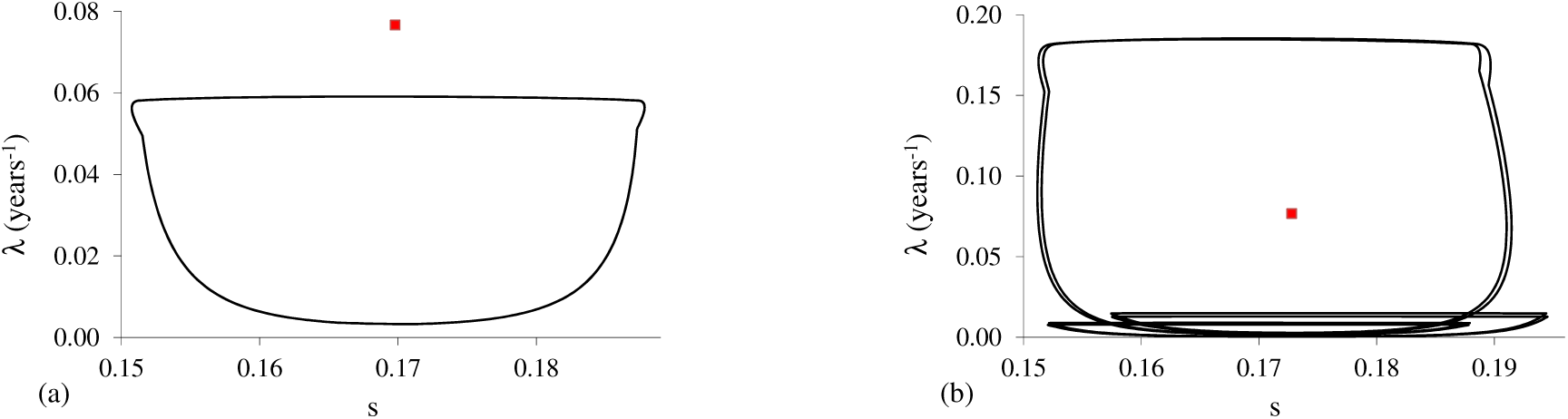
Asymptotic curves for: (a) *p* = 18.972%, one cycle orbit, and (b) *p* = 18.974%, six cycles orbit.

In Figure 9(b), when *p* = 17.437%, a one cycle orbit contains *P*^*^ quasi at the boundary. The minimum value of the force of infection, *λ* = 1.6 × 10^−5^ *years*^−1^, occurs at *t* = 4.9 *years*, or, *i* = 9 × 10^−8^ (90 individuals in 1 billion). When *p* = 18.972%, a one cycle orbit does not contain *P*^*^ anymore, and the minimum value of the force of infection, *λ* = 1.8 × 10^−11^ *years*^−1^, occurs at *t* = 153.2 *years*, or, *i* = 1 × 10^−13^ (10 individuals in 1 trillion). For *p* = 18.975%, one cycle orbit appears again, which is very similar than that observed in Figure 11(a). The period of closed orbit is *T* = *ετ*, which persists until *p* = 18.977%. However, for *p* = 18.978%, three cycles orbit arises, which is shown in Figure 12: (a) initial time (until 50 *years*), and (b) asymptotic curve. The period of closed orbit is *T* = 3*ετ*, persisting until *p* = 18.978%, and at *p* = 18.979% one cycle orbit arises.

**Figure 12:**
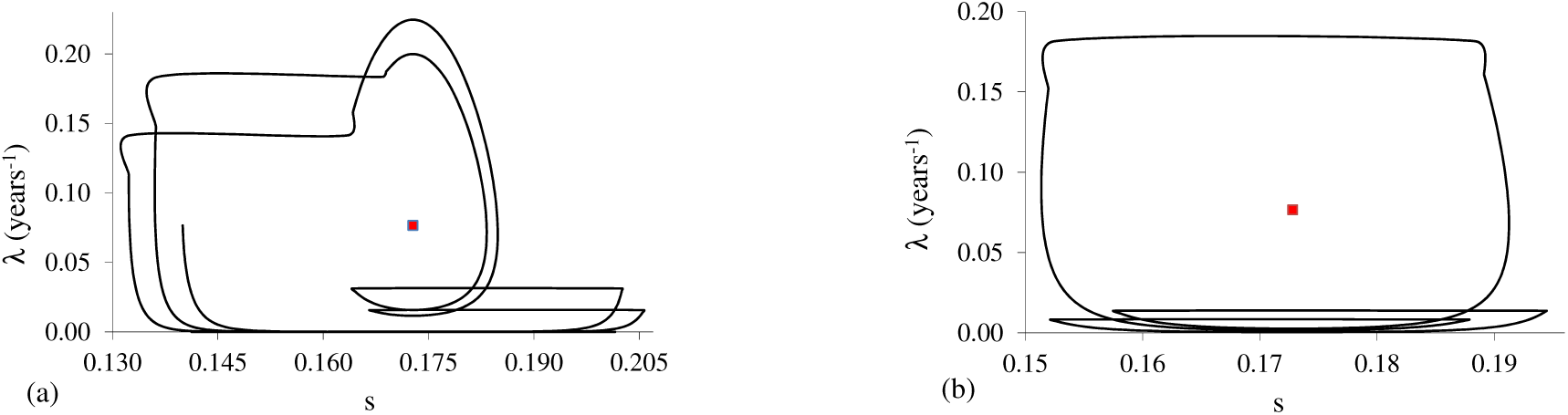
Phase space portrait for *p* = 18.978%: (a) initial time (until 50 *years*) and (b) asymptotic curve, a three cycles orbit.

There are alternating one cycle – three cycles orbits (60 alterations were observed between 18.979% and 19.367%). Again a one cycle orbit does not contain *P*^*^. Additionally, three special trajectories are found in this interval. (1) At *p* = 19.010% there are not closed orbits, but the trajectories are bounded in a strip for small error *ϵ* ≤ 8.0 × 10^−5^. Figure 13(a) shows the asymptotic curve. However, for lower accuracy (the error is increased, *ϵ* ≥ 8.1 × 10^−5^), then *T* = 72*ετ* = 237.1968 *years*, that is, the period is increased in 72 times. (2) Figure 13(b) shows the asymptotic curve for the case *p* = 19.018%. The period of closed orbit is *T* = 18*ετ* = 59.2992 *years*, which persists until *p* = 19.019%.

**Figure 13:**
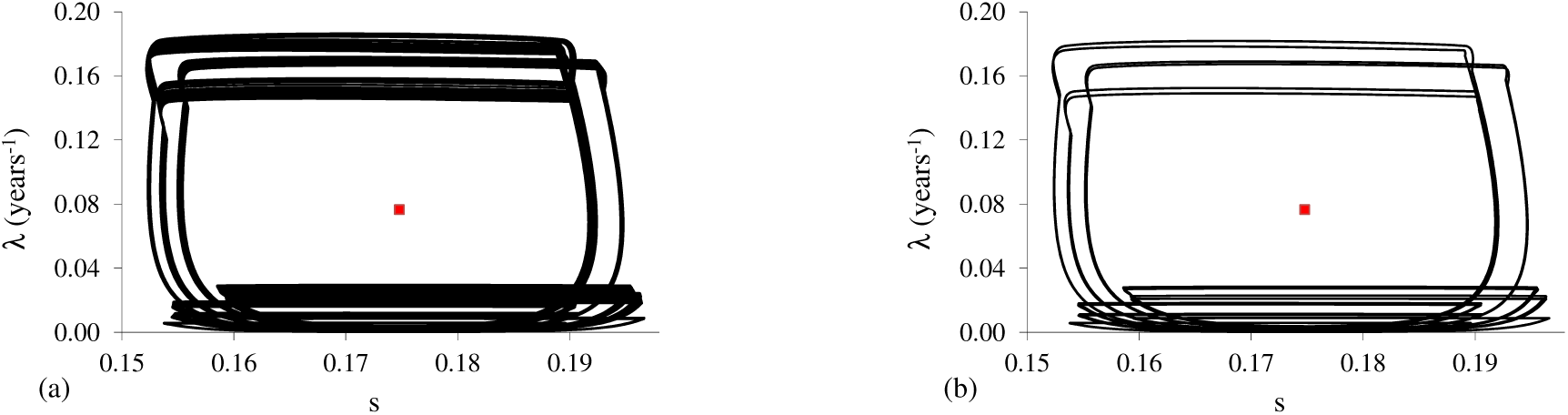
Asymptotic curve for: (a) *p* = 19.010%, irregular but a seventy two cycles orbit for decreased relative error *ϵ*, and (b) *p* = 19.018%, an eighteen cycles orbit.

(3) The asymptotic curve for the case *p* = 19.022% is similar to figure shown in Figure 13(b), but the period of closed orbit is *T* = 9*ετ* = 29.6496 *years*, which occurs again at *p* = 19.029%.

With respect to the alternating one cycle – three cycles orbits, the last three cycles orbit occurs between 19.366% and 19.367%. At *p* = 19.368% one cycle orbit is observed, and the minimum value of the force of infection, *λ* = 8.1 × 10^−13^ *years*^−1^, occurs at *t* = 169.5 *years*, or, *i* = 4.6 × 10^−15^ (5 individuals in 1 quadrillion). Figure 14 shows the asymptotic curves for: (a) *p* = 19.367% (*T* = 3*ετ*), and (b) *p* = 19.368% (*T* = *ετ*).

**Figure 14:**
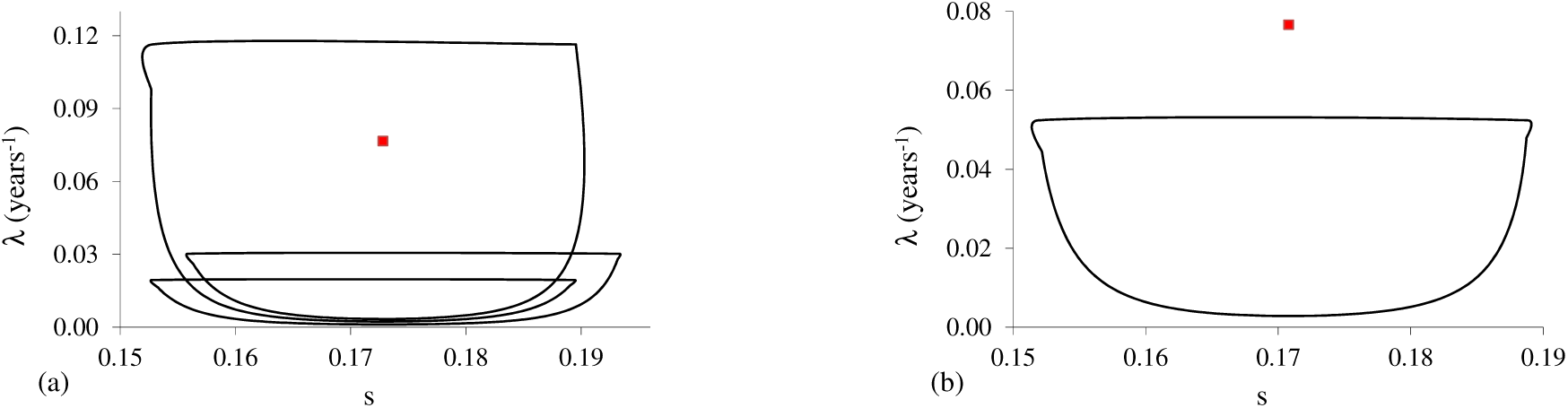
Asymptotic curves for: (a) *p* = 19.367%, a three cycles orbit, and (b) *p* = 19.368%, a one cycle orbit.

After *p* = 19.368% only one cycle orbit is observed, which persists until *p* = 22%. Figure 15 shows for *p* = 22%: (a) initial time (until 50 *years*), and (b) long time (from 400 to 800 *years*). The period of closed orbit is *T* = *ετ*. The force of infection decreases up to 10^−15^ *years*^−1^(*i* = 5.6 × 10^−18^), but the maximum value of *s* is near 0.193, which is higher than *s*_0_ = 0.1728 (Figure 15(a)). For this reason the force of infection increases after 65 *years*, and approaches to 0.008 *years*^−1^ (Figure 15(b)).

**Figure 15:**
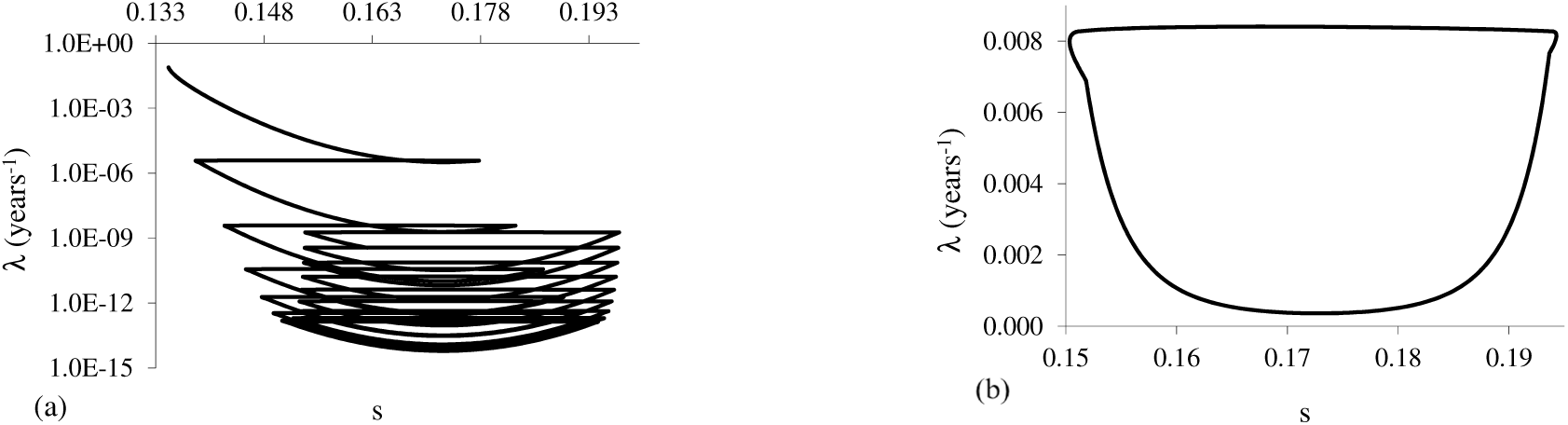
Phase space portrait for *p* = 22%: (a) initial time (until 50 *years*) and (b) long time (from 400 to 800 *years*), an one cycle orbit.

At and after *p* = 23% there are not closed orbits: the fraction of susceptibles oscillates, but the force of infection always decreases. However, these oscillations have two behaviors. One of them is the maximum value of susceptibles being higher than the equilibrium value *s*_0_. In the second type, the maximum value of oscillating *s* situates always lower than *s*_0_. Figure 16 shows the phase space portrait (from 0 to 50 *years*) for: (a) *p* = 23%, and (b) *p* = 27%. Notice that Figure 16(a) is quasi similar than that shown in Figure 15(a), but asymptotically there is not closed orbit for the force of infection (*λ* is always decreasing).

**Figure 16:**
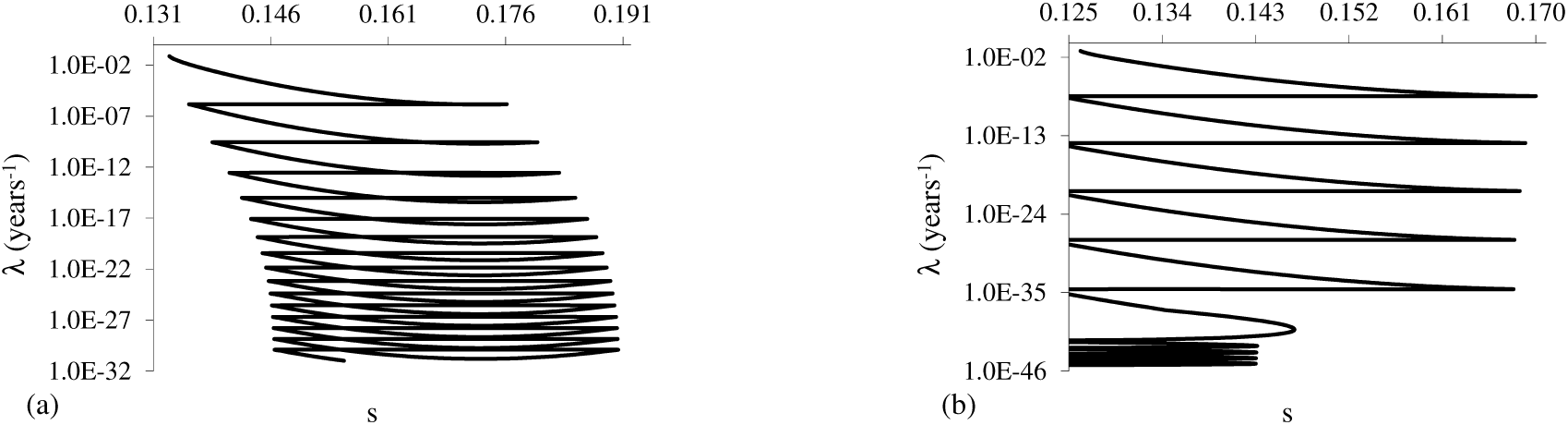
Phase space portrait (from 0 to 50 *years*) for: (a) *p* = 23%, with *s* oscillating beyond the equilibrium value, and (b) *p* = 27%, with *s* oscillating below the equilibrium value.

When a vaccination does not eradicated the disease, there is a well known result about the fraction of susceptible individuals: the steady state fraction of susceptible individuals just before the introduction of the vaccination is equal to the asymptotic fraction of susceptible individuals after vaccination [2] [24]. In above simulations dealing with a series of pulse vaccinations, for *p* ≤ 23%, the fraction of susceptible individuals oscillates above the equilibrium value *s*_0_. However, for *p >* 23%, the fraction of susceptibles does not surpass *s*_0_ anymore, rather the successive peaks decrease assuming values lower than *s*_0_. Hence, *p* near 23% can be assumed as the proportion of susceptibles to be vaccinated to eradicate the disease in the dynamical point of view.

## 4 Discussion

In this paper, neither the vertical transmission nor vaccination of newborns were considered. The proportion of susceptibles being vaccinated *p* was varied broadly, fixing the period at *T* = *ετ*. Let the variation of susceptibles and periodic orbits be discussed.

In all simulations, the range of variation of the fraction of susceptible individuals are narrow in comparison with the variations in the force of infection. The largest amplitude of the variation of the fraction os susceptible individuals divided by the equilibrium fraction *s*_0_ is around (0.22 − 0.13) */*0.173 ∼ 0.52 at *p* = 15.22%; however, for the force of infection, this quotient is 0.6*/*0.078 ∼ 7.69 at *p* = 15.71%. This fact shows the necessity of choosing carefully the proportion of susceptibles to be vaccinated (the risk of a severe epidemics is higher for *p* near 15%, which is in the range 8.987% *< p <* 19.368% where complex behavior occurs). All the foregoing scenarios show that, when the disease is not eradicated, the possible outbreak of an epidemics is not negligible, especially in the scenarios where closed orbits with multiplicity of basic period *T* occur.

There are clearly two windows in the proportion vaccinated *p* at which a single closed orbit occurs with period *T* = 3.2944 *years*: 0 *< p* ≤ 8.987% and 19.368% ≤ *p <* 22%. Between 8.987% *< p <* 19.368% (excluding 17.437% *< p <* 18.926% where single orbit occurs), closed orbits with multiplicity of basic period *T* are observed, besides bounded trajectories (torus bifurcation). After *p >* 22%, none periodic orbits are observed. When 22% *< p* ≤ 23%, the non-periodic trajectories oscillates beyond the value *s*_0_; but for *p >* 23%, the oscillations occur below this value. Notice that one cycle is observed in the range 19.368% ≤ *p <* 22%, but this one-cycle arises after transient behavior where the number of infectious individuals reaches extremely small value, for instance, *i* = 10^−18^ (see Figure 15). Notably, at *p* = 18.972% and above this value, when a one cycle orbit occurs, this closed orbit does not contain *P*^*^ anymore, and the minimum value for the infectious individuals is very small, for instance, 10 individuals in 1 trillion for *p* = 18.972%.

In order to understand the breaking of cyclic regime (*p >* 22%), let a constant and continuous vaccination scheme be recalled. In this modeling, the asymptotic fraction of susceptible individuals always reaches the same value 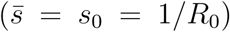 regardless the vaccination rate, if this rate is lower than the threshold *ν*^*th*^ given by equation (5). However, if the vaccination rate is higher than the threshold (*ν > ν*^*th*^), the asymptotic fraction of susceptible individuals attains 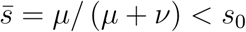, which is obtained by letting *λ* = 0 in equation (2). Following this reasoning, the proportion *p* at which the fraction of susceptible individuals oscillates below the value *s*_0_ could be considered as the threshold of proportion vaccinated *p*_min_. In this case, the minimum proportion to be vaccinated is *p*_min_ = 23% in order to achieve eradication.

However, numerical simulations showed the possibility of choosing two other proportions of vaccination in which the disease can be considered eradicated. The first is the disappearance of cyclic behavior, which occurs when *p >* 22%, near to the *p*_min_ = 23% obtained previously. The second is the appearance of the first single orbit where the equilibrium point is outside the cycle, which occurs at *p* = 18.972%. In both cases, the number of infectious individuals achieves extremely low value.

The occurrence of periodic solutions is compared with a simple discrete model, and the results from sinusoidal modeling are discussed.

### 4.1 Bifurcation in discrete model

May [13] showed, dealing with a discrete logistic map

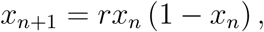

where *r* is the birth rate and *x*_*n*_ is the fraction of a population in *n*-*th* generation, the appearing of 2^*k*^-cycle as *r* increases. (Notice that the discrete size of population is *X*_*n*_ = *Kx*_*n*_, where *K* is the carrying capacity.) Further increase in *r* results in the appearing of odd cycles: 3-cycle, followed by 6-cycle and further period doubling. For higher *r*, chaos is found. When period doubling occurs, there are attractors with small and high values, which difference increases with increasing *r*.

The first transcritical bifurcation, when there is a doubling of period from one cycle, is explained as follows. As the birth rate *r* increases, the number of individuals increases, but reaching a certain value, the corresponding fraction of population destabilizes by the fact that it cannot bear such amount anymore. Hence, after the first bifurcation value of *r* occurs a jump between two values, that is, one lower and other higher than the corresponding unstable value, and the difference between these values (fixed points) increases as *r* increases. The biological meaning of this first doubling of period is the role played between food availability and replenishing of population: a high size of population exhausts the food, and the next generation is settle at a lower size; which in turn allows elevate number in the next generation due to abundance of food. The second transcritical bifurcation destabilizes these two fixed points, allowing the appearance of four fixed points. The reason is that the depression of next generation due to the highest level of population is so great that two intermediate levels (fixed points) are needed as steps to return to the highest level. This explanation follows for higher number of fixed points.

However, in the corresponding continuous modelling, the logistic equation *dX/dt* = *rX* (1 − *X/K*), where *K* is the carrying capacity and *X* is a continuous size of a population, there is a single closed orbit with period given by the time lag between successive impulses [6]. In this particular example (the solution *X*(*t*) is monotonically increasing), there are not transient trajectories. However, in pulse vaccinations modelling, closed orbits arise after a period of time when transient dynamics fades out, and this transient period is increased as the perturbation (proportion vaccinated *p*) increases. In some cases, the transient dynamics persists indefinitely, in which case closed orbit does not occur.

In some extent, those results found in [13] can explain the bifurcations occurred in pulse vaccination strategy analyzed here. (In a pulse vaccination modelling, the accumulation of susceptible individuals by birth plays the role of limited food and space in the logistic map.) In another words, if the vaccination (perturbation of the system) *p* increases, then a huge epidemics exhausts susceptible individuals, and during the inter-vaccination period occurs a small accumulation of susceptibles, triggering next mild epidemics. However, this mild epidemics left relatively high size of susceptible individuals, which, gathered with the accumulated susceptibles during inter-vaccination period, originates huge epidemics, and so on.

### 4.2 Sinusoidal contact rate modeling

Now, let a forced SIR model be considered. Instead of pulses of vaccination, the model described by equation (7) takes into account the periodicity in the transmission rate, mimicking a seasonal variation during a year. This model is described by

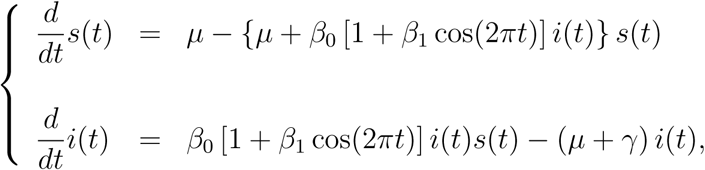

where the transmission coefficient *β*(*t*) is an annual periodic function with *t* in *years*. By varying *β*_0_ (*β*_1_ is fixed in 0.28) from 500 to 1800 *years*^−1^, Rand and Wilson [20] obtained stability of annual limit cycle, but for higher values, they observed biennial cycle, which was followed by period doubles and then chaotic attractor. They observed very long-term transient dynamics. The origin of the period doublings is again the amount of susceptible individuals. As the logistic map where limited space and food (characterized by a constant carrying capacity *K*) disturbs the size of population for large birth rate *r*, the accumulation of susceptible individuals between successive outbreaks results in period doublings: small epidemics is succeeded by large epidemics, as discussed above.

When periodically forced systems are considered, there are two main features: (1) transient dynamics, which lasts for long time as the amplitude of the forced input increases, and (2) the period doubling bifurcation, eventually arising irregular and chaotic behaviors. However, higher order periodic cycle and even chaotic behavior occurs when the parameter *β*_0_ assumes higher values, such that *i <* 10^−10^ [20], that is, less than 1 infectious in 10 billions.

## 5 Conclusion

In the literature, there are many papers dealing with pulse vaccination strategies. Jiang and Yang [11] analyzed SIR model with birth pulse and pulse vaccination. By fixing the proportion of susceptibles being vaccinated, they varied the maximum birth rate, and they constructed a bifurcation diagram similar to that obtained by May [13]. Meng and Chen [14] considered SIR model encompassing vertical transmission also, and studied pulse vaccination applied on newborns. They established conditions for the stability of infection-free periodic solution. Nie *et al*. [15] considered SIR model with state dependent pulse vaccination. They proved the existence and stability of positive order-1 and order-2 periodic solutions. Qin *et al*. [19] varied the time between successive pulses vaccinations in SIR model, and obtained a bifurcation diagram. Indeed, Qin *et al*. [19] considered the varying proportion of vaccination of susceptibles be dependent on susceptible individuals and on the resource limitation parameter.

In this paper, a pulse vaccination modeling was dealt numerically in order to understand the origin of cycles in a periodically forced systems. Dynamical trajectories were shown instead of bifurcation diagram using Poincaré map [20]. A simple periodic impulsive dynamical system analyzed here presented period doublings bifurcation, and very long lasting transient, which is essentially a irregular trajectory. These two characteristics are found in periodically forced dynamical system, for instance in sinusoidal contact rate. The accumulation of susceptible individuals in forced epidemiological modeling plays the role of constant carrying capacity in a discrete one dimensional logistic map. In both cases, when doubling of periods occurs, there arise small and large orbits (stable fixed points) from destabilizing orbit (unstable point).

The arising of long term complex dynamics (higher order periodic cycles and irregular behaviors asymptotically) occurs in a range of parameters leading to extremely small number of infectious individuals (for instance, less than one in 10 billions). Also, the transient behavior lasts for a long time. For this reason, in mathematical modelling with periodic external forces, the existence of closed orbits and chaos is nice mathematical results, but appears to be meaningless biologically. For instance, the knowledge of the closed orbits arising after hundreds of years seems not be relevant to the public health authorities; on the contrary, irregular transient dynamics must be taken into account, where huge epidemics can occur occasionally. Hence, due to the long lasting transient behavior, the description of biological phenomena by mathematical modelling should be restricted in a pre-determined period of time. Also, experimentally observed time varying model parameters should be preferable instead of well behaved continuous periodic functions. For instance, Yang *et al*. [25] considered time varying temperature and rainfall in the model parameters.

## Data Availability

All data produced in the present work are contained in the manuscript

